# Treatment Outcomes for Adult Patients with Localized Osteosarcoma Treated with Chemotherapy without Methotrexate

**DOI:** 10.1101/2020.05.27.20112730

**Authors:** MRM Silva, RC Bonadio, GDR Matos, D Toloi, M Nardo, RM Munhoz, CLCB Zampieri, Daniel Reboledo, LFM Correa, AM Baptista, O Feher, VP Camargo

## Abstract

**Background:** Standard treatment for pediatric patients with localized osteosarcoma include high-dose methotrexate (HDMTX) and cure rates greater than 60% are observed. In adult patients, however, the toxicity profile limits the use of HDMTX and it is usually excluded from chemotherapy protocols for this group. We aimed to evaluate the outcomes of adult patients with localized osteosarcoma treated with chemotherapy without methotrexate.

**Methods:** In this retrospective cohort, we evaluated adult patients with high-grade osteosarcoma who received treatment with chemotherapy without methotrexate in a reference cancer center from 2007 to 2018. Outcomes analyzed were recurrence-free survival (RFS), overall survival (OS), and prognostic factors associated with overall survival.

**Results:** A total of 48 patients had localized disease and received treatment with chemotherapy without methotrexate. The majority of them received chemotherapy with a combination of cisplatin and doxorubicin (N=42, 87.5%). Median age was 27 years. (range 16.8 – 66.7) With a median follow-up of 29.2 months, median RFS was 29.9 months. Median OS was not reached. 5-year RFS and OS rates were 35.1% (95% CI 20.3 – 50.2%) and 71.6% (95% CI 52.3 – 84.2%), respectively. Patients who received cumulative doses of doxorubicin ≥ 375 mg/m2 had better OS than those who received lower doses (HR 0.26, 95% CI 0.07 – 0.94, P = 0.041). Similarly, patients who received ≥ 6 cycles of neoadjuvant/ adjuvant cisplatin tended to have better OS than those who received < 6 cycles (HR 0.30, 95% CI 0.08 – 1.09, P = 0.069).Nineteen patients received less than 6 cycles of cisplatin and doxorubicin mainly because of grade 3 or 4 toxicities (11), disease progression (6),patient refusal(1), physician choice(1).

**Conclusion:** In our study, adult patients with localized high-grade osteosarcoma who were treated with chemotherapy without methotrexate had unfavorable outcomes. The cumulative doxorubicin dose and the number of cisplatin/doxorubicin cycles were associated with improved OS. The investigation of additional treatment strategies is of utmost importance to improve adult patients outcomes.

## Introduction

Osteosarcoma is a rare disease in adults, and is the most common primary bone malignancy in children and young adolescents.^1^ Most patients present localized disease^2,3^ and cure rates are around 60-70% in the pediatric population with contemporary treatments regimens. ^4,5,6,7^. These results were established by several landmark studies during 1970s and 1980s. They demonstrated the efficacy of adjuvant chemotherapy and later neoadjuvant chemotherapy.^8^ In a large Brazilian study published in 2013 with 390 non metastatic pediatric and young adults osteosarcoma patients, the median survival and relapse free survival in 5 years were 59% and 48%, respectively. This is the largest study with osteosarcoma patients in a developing country reflecting more the reality of our cohort.

In the pediatric population, the treatment of localized osteosarcoma consists of a multi-agent chemotherapy with usually includes high doses of methotrexate, doxorubicin, and cisplatin.^5,6,7^ However, in adults, several studies questioned the value of methotrexate, due to the greater risk of serious toxicities like nephrotoxicity, otoxicity, mucositis, hepatotoxicity, pulmonary toxicity, and neurotoxicity (1).^9^ As in other prospective studies, the impact of adding methotrexate to the chemotherapy regimen failed to demonstrate an increase in overall survival. ^10,11,12,13^

Considering this, standard chemotherapy for adult patients frequently does not include high-dose methotrexate. A comprehension of the outcomes of these patients without methotrexate is important to understand the disease scenario of this selected population and to guide the development of new treatment strategies with acceptable tolerance. The present study aimed to evaluate the outcomes of adult patients with high-grade localized osteosarcoma treated with chemotherapy regimens without methotrexate.

## Methods

### Study design and Participants

In this single-center retrospective cohort, adult patients with high-grade osteosarcoma who received treatment at a Brazilian tertiary cancer center *(Instituto do Câncer do Estado de São Paulo*, São Paulo, Brazil) between 2007 and 2018 were evaluated. Electronic medical records were reviewed to collect patients’ data, including clinical and demographical characteristics, treatment received, and outcomes.

Patients were included in the analysis if they presented with histologically confirmed localized high-grade osteosarcoma and received neoadjuvant, adjuvant or perioperative treatment with a chemotherapy regimen that did not include methotrexate. Additionally, patients should be at least 16 years old. Exclusion criteria were the presence of distant metastasis, the presence of localized disease not amenable to curative surgery at diagnosis, and diagnosis of a secondary malignant neoplasm in the past 5 years (except for non-melanoma skin cancer).

Local Ethics Committees approved the present study.

### Treatment

Systemic treatment consisted of neoadjuvant or adjuvant chemotherapy with regimens based on combinations of cisplatin, doxorubicin, ifosfamide, and/ or etoposide. Perioperative chemotherapy with chemotherapy cycles both pre and post-surgery was also acceptable. The timing of chemotherapy in relation to surgery was defined by physicians’ discretion.

All patients had curative surgery planned as part of their oncologic treatment. The planned surgery was not performed only in case of patients’ refusal, disease progression during neoadjuvant chemotherapy or other complication that contraindicated surgery.

### Statistical analysis

Patients and treatment characteristics were summarized using descriptive statistics. Median and range were used to summarize continuous variables. Categorical variables were presented using their absolute and relative frequencies.

The outcomes evaluated were overall survival (OS) and recurrence-free survival (RFS). OS was defined as the time from diagnosis until death from any cause. RFS was the time from diagnosis until disease recurrence or death, whichever occurred first. Patients without these events were censored at the time of last follow-up.

Survival analyses were performed using the Kaplan-Meier method, with the log-rank test to compare the difference between survival curves when appropriate. Factors associated with OS were investigated using univariate Cox proportional hazards model.

Factors included in the univariate analysis were: age (≥ 40y vs < 40y), ECOG-performance status (≥ 2 vs 0-1), Huvos criteria after neoadjuvant chemotherapy (3-4 vs 1-2), number of cisplatin cycles (≥ 6 vs < 6 cycles), cumulative dose of doxorubicin (≥ 375 mg/m^2^ vs < 375 mg/m^2^), occurrence of grade 3-4 toxicities (yes vs no), and type of surgery (amputation vs preservation) The Huvos a classification is a grading system from I-IV of the pathological response after chemotherapy, with higher grades being associated with better treatment response (2).

For statistical significance, we considered P values less than 0.05. Statistical analyses were performed using Stata software, version 15.1 (StataCorp, Texas, USA).

## Results

### Patients’ characteristics

A total of 97 adult patients with osteosarcoma were treated at ICESP from 2007 to 2018. Sixty-one patients had localized resectable disease at diagnosis and 48 of them (78.7%) received neoadjuvant or adjuvant chemotherapy without methotrexate, composing the study population.

Median age was 27 years (range 16.8 – 66.7) and the majority of patients (93.7%) presented primary tumor site at extremities. The most common histologic subtype was osteoblastic (43.7%). Patients’ clinical and demographical characteristics are summarized in **Table 1**.

**Table 1.** Patients’ characteristics

|  | No of patients<br>(N = 48) | % |
| --- | --- | --- |
| Age, years |  |  |
| Median (range) | 27 (16.8 – 66.7) |  |
| Gender |  |  |
| Male | 28 | 58.3 |
| Female | 20 | 41.7 |
| ECOG-PS |  |  |
| 0 | 6 | 12.5 |
| 1 | 29 | 60.4 |
| 2 | 12 | 25 |
| 3 | 1 | 2.1 |
| Primary tumor site |  |  |
| Extremity | 45 | 93.7 |
| Axial | 3 | 6.2 |
| Initial TNM stage |  |  |
| I | 3 | 6.2 |
| II | 34 | 70.8 |
| III | 9 | 18.7 |
| NA | 2 | 4.2 |
| Histologic type |  |  |
| Osteoblastic | 21 | 43.7 |
| Condroblastic | 4 | 8.3 |
| Mixed | 8 | 16.7 |
| Pleomorphic sarcoma | 3 | 6.2 |
| Others | 10 | 20.8 |
| NA | 2 | 4.2 |
| Chemotherapy timing |  |  |
| Neoadjuvant | 10 | 20.8 |
| Adjuvant | 10 | 20.8 |
| Perioperative (Neoadjuvant + Adjuvant) | 28 | 58.4 |
| Huvos* |  |  |
| I | 15 | 39.5 |
| II | 9 | 23.6 |
| III | 7 | 18.4 |
| IV | 1 | 2.6 |
| NA | 6 | 15.8 |
| Surgery type |  |  |
| Limb preservation | 29 | 60.4 |
| Amputation | 19 | 39.6 |

Abbreviations: ECOG-PS, Eastern Cooperative Oncology Group Performance Status; TNM, tumor, node, metastasis; NA, not available.
\* Huvos criteria for patients who were submitted to neoadjuvant chemotherapy (N = 38).

The chemotherapy regimen received by most of the patients was a combination of cisplatin and doxorubicin (N=42; 87.5%). The other six patients received cisplatin, doxorubicin, ifosfamide, and etoposide (N=6; 12.5%). Median number of cisplatin/doxorubicin cycles was 6 (range 3 – 8) and median cumulative dose of doxorubicin was 375 mg/m^2^ (range 225 – 495 mg/m^2^).

### Patients’ outcomes

Patients were followed for a median time of 29.2 months. During the follow-up period, 27 patients had disease recurrence or death. Median RFS was 29.9 months (95% CI 11.3 – NR months). Five-year RFS rate was 35.1% (95% CI 20.3 – 50.2%). Ten of the 48 patients died, with the median OS not reached. Five-year OS rate was 71.6% (95% CI 52.3 – 84.2%). Kaplan-Meier survival curves are shown in **Figure 1**.

**Figure 1.**
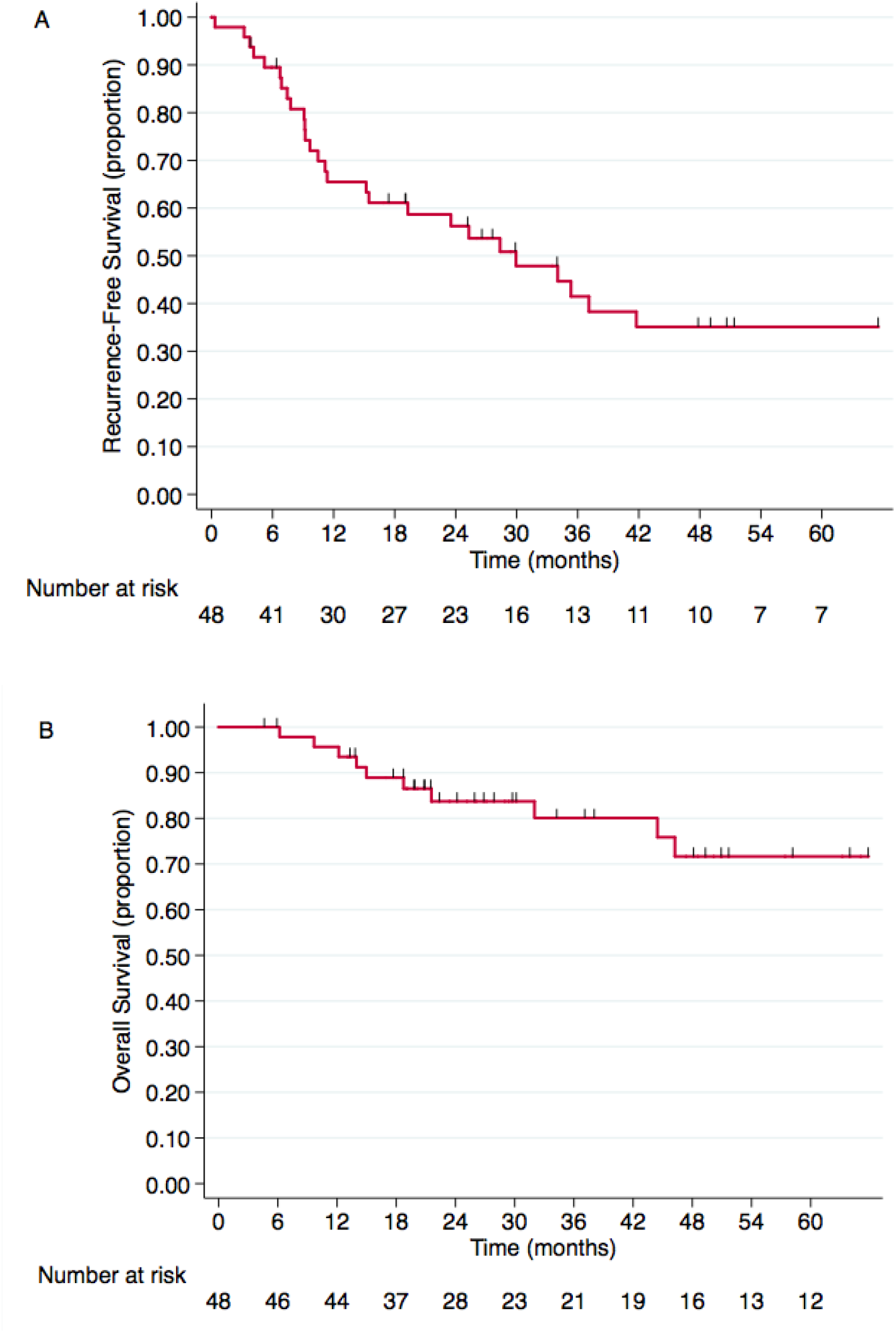
Kaplan-Meier curves for recurrence-free survival (a) and overall survival (b) of adult patients with localized high-grade osteosarcoma treated with chemotherapy without methotrexate.

In the univariate analysis, higher cumulative doses of doxorubicin (≥ 375 mg/m^2^) were associated with improved OS in comparison with lower doses (< 375 mg/m^2^) (HR 0.26, 95% CI 0.07 - 0.94, P = 0.041). Patients who received a cumulative doxorubicin dose more or equal to 375 mg/m^2^ had a 5-year OS rate of 79.5% compared to 45% among those who received lower doses (P log-rank = 0.029). In addition, another factor that tended to be associated with OS was the number of cisplatin/doxorubicin cycles (neoadjuvant and adjuvant) (HR 0.30, 95% CI 0.08 – 1.09, P = 0.069). The results of the univariate analysis of factors associated with overall survival are presented in **Table 2**. **Figure 2** shows the Kaplan-Meier curves for OS according to the cumulative dose of doxorubicin received.

**Table 2.**
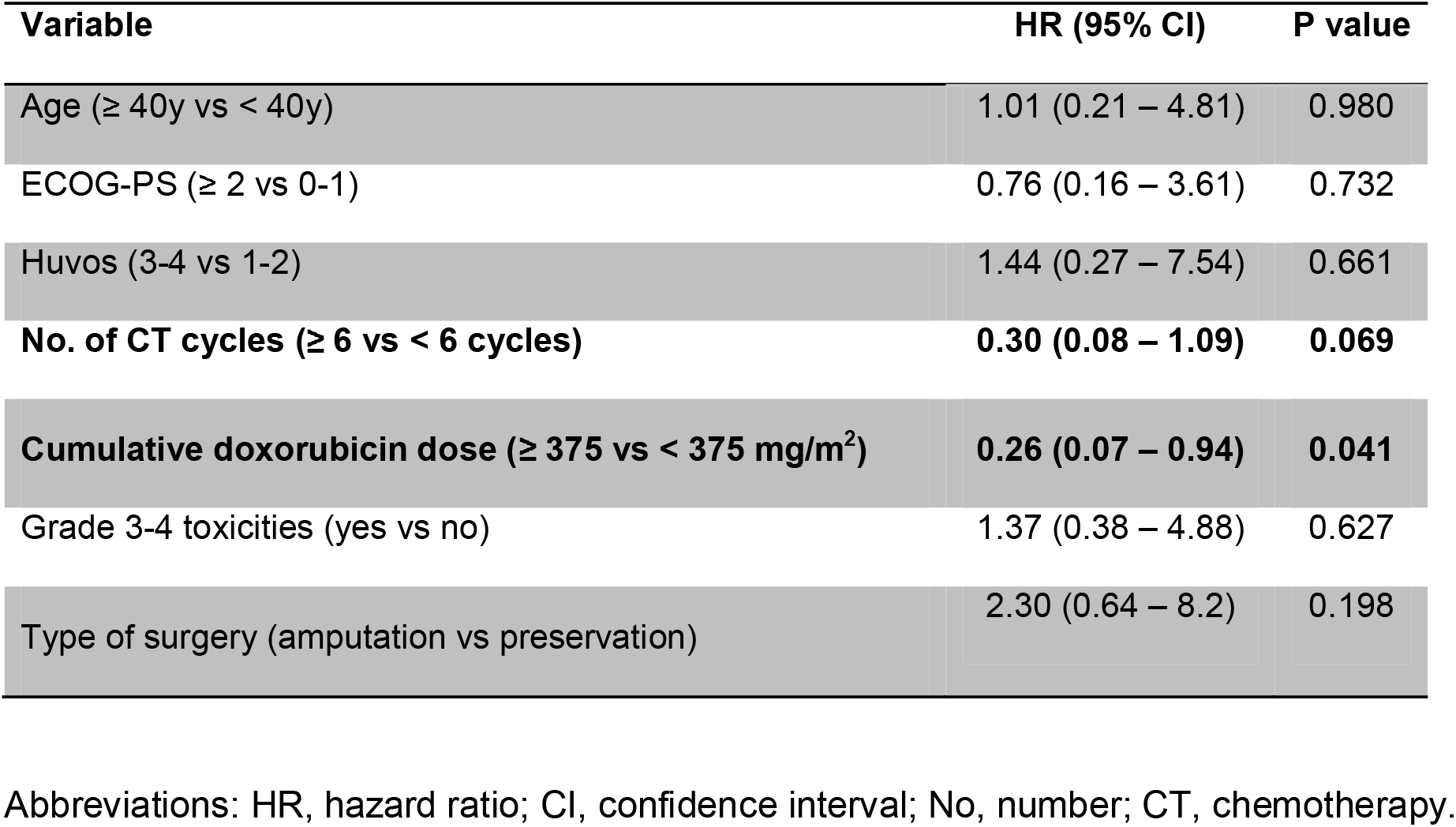
Factors associated with overall survival (Cox proportional hazards model).

**Figure 2.**
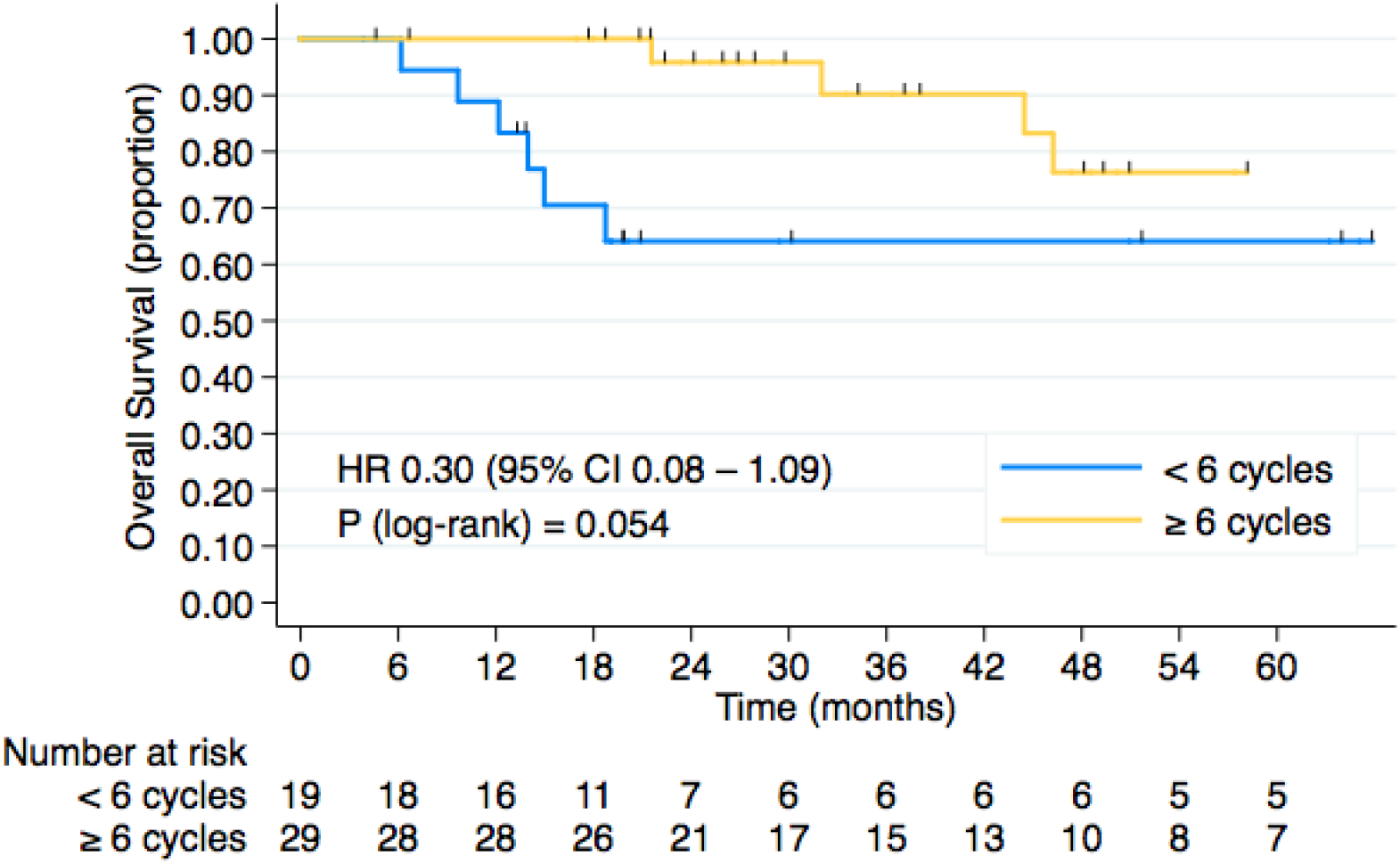
Kaplan-Meier curves for overall survival of adult patients with localized high-grade osteosarcoma treated with chemotherapy without methotrexate, according to cumulative dose of doxorubicin received (neoadjuvant, adjuvant or perioperative).

Main reasons for receiving less than six cycles were grade 3-4 toxicities (N=11/19, 57.9%) and disease progression during chemotherapy (N=6/19, 31.6%). In the overall study population, 17 patients (35.4%) had grade 3-4 toxicities.

## Discussion

In the present study, adult patients with localized osteosarcoma treated with chemotherapy regimens without high-dose methotrexate had unfavorable outcomes, with only 35% disease-free in 5 years and an overall survival of 71,5%. The only factor associated with overall survival was the chemotherapy dose received, with a cumulative doxorubicin dose greater than 375 mg/m^2^ and the number of cisplatin/doxorubicin cycles greater than 6. Similarly, in the non metastatic cohort of the largest Brazilian pediatric study with a median follow up of 92 months, the 5 year overall survival and relapse free survival was 59% and 48%, respectively. The most important factors associated with survival were: metastases at diagnosis, grade of necrosis and type of surgery. Both cohorts included patients with locally advanced disease at diagnosis with amputation rates around 40% in both cohorts, probably related to the difficulty for this patients to reach a specialized sarcoma center in Brazil.^3^

In a study by Souhami et al., 407 patients with non-metastatic osteosarcoma were randomized to receive cisplatin and doxorubicin or vincristine, high-dose methotrexate, and doxorubicin in the preoperative setting or bleomycin, cyclophosphamide, actinomycin D, vincristine, high-dose methotrexate, doxorubicin, and cisplatin in the postoperative setting (3). Only 51% of patients were able to complete treatment with multiple drugs versus 94% with the two-drug regimen. The percentage of patients with tumor necrosis greater than 90% was 29% in both groups. In addition, the 5-year overall survival rate was 55% in both. This study was criticized for the low overall survival in the high-dose methotrexate group, compared to previous studies^4^. In another randomized study, the addition of methotrexate to cisplatin and doxorubicin regimen resulted in lower disease-free survival (DFS) (5-year DRS rate: 57% versus 41%, P = 0.02) and overall survival (5-year OS rate: 64% versus 50%, P = 0.10) (4). In a subgroup of patients with necrosis greater than 90% (N=66), greater progression-free and overall survival was observed.^5^

Another recent study by Wippel B et al, evaluated 33 pediatric and adult patients with localized osteosarcoma submitted to high-dose methotrexate (12 g/m^2^) with cisplatin and doxorubicin^6^. They found a statistically significant delay in clearance of methotrexate (120vs 79 hours, P < 0.001) as well as a lower number of cycles in the adult patients (12 vs 5 cycles, P < 0.01). The authors explained that subsequent delays in doxorubicin/cisplatin therapy due to slow methotrexate clearance may arguably have a negative impact on disease outcomes as well and may play a role in physician decision making process in these situations. Histologic response is a well-established prognostic indicator in osteosarcoma, and, in this study, histologic response was positively correlated with the number of preoperative high-dose methotrexate doses received. A trend toward improved survival was noted for patients who received at least seven doses of high-dose methotrexate^6^. However, in our study, we found that the number of cisplatin and doxorubicin cycles correlated with outcome.

In addition, a recent Brazilian study from our group performed a retrospective study of 10 patients aged 16-23 years who received high-dose methotrexate. Two of them died of therapy related causes^7^. The EUROpean Bone Over 40 Sarcoma Study (EURO BOSS) prospectively gathered data on high-grade osteosarcoma patients ages 41‐65, an older population than studied here. EURO-BOSS patients received investigator’s choice therapy and included 48 patients who received high-dose methotrexate. Among these older patients, 23% experienced delayed methotrexate clearance and only one developed ≥ grade 2 nephrotoxicity.^8^

In our center, the clearance delay of methotrexate together with the high toxicity in the adult population and the loss of dose intensity of cisplatin and doxorubicin contributed to exclude the drug from our treatment protocol in adult osteosarcoma. In face of this, another treatment strategy that we studied was the addition of ifosfamide and etoposide to the adjuvant treatment.

We included six patients in a phase II study evaluating the addition of 6 cycles of ifosfamide and etoposide (IE) after surgery for patients who have already received up to 6 cycles of cisplatin and doxorrubicin pre operativel.^8^ We compared this group to another 6 matched patients that received 3 cycles of Cisplatin and Doxorubicin (standard group) before and after surgery. Median age was 20.1 years. Grade ≥ 3 toxicities occurred in 83% of the patients in IE group, including one grade 5 toxicity. The study was interrupted because of the alarming toxicity. In addition, numerally lower recurrence-free survival (median: 11.3 vs 31.8 months; HR 1.68, 95% CI 0.36 - 7.89) and overall survival rates (median: 44.4 vs 51.2 months; HR 1.57, 95% CI 0.29 - 8.46) were observed in IE group.^9^ Thus, studying additional treatment strategies is still warranted to improve outcomes of adult patients.

In conclusion, adult osteosarcoma remains a challenge disease with most treatments based on pediatric studies. In the present study, patients with localized osteosarcoma treated with chemotherapy without methotrexate had unfavorable outcomes in comparison with the worldwide pediatric population but similar to the Brazilian pediatric population.In developing countries, patients arrive to reference centers with locally advanced disease what justifies the bad outcomes in both pediatric and adult population.

Nevertheless, the use of high-dose methotrexate (12 g/m^2^) remains controversial in adult patients and, in our opinion should be used, only inside clinical trial protocols. The clearance delay of methotrexate may contribute to postpone the subsequent cycles of the main drugs cisplatin and doxorubicin, interfering directly in the final outcome.In our study the number of cycles of cisplatin and doxorubicin as well as the accumulative doxorubicin dose was correlated to better survival. However, 35% of the patients had a dose reduction because of grade 3 or 4 toxicity even without Metotrexate. We definitively need more multi-institutional studies to elaborate an adult osteosarcoma guideline.

## Data Availability

Enclosed is a manuscript entitled Treatment Outcomes for Adult Patients with Localized Osteosarcoma Treated with Chemotherapy without Methotrexate to be considered for publication in Esmo Open.
Osteosarcoma is a rare disease in adults, and is the most common primary bone malignancy in children and young adolescents. Most patients present localized disease and cure rates are around 60% in the pediatric population with contemporary treatments regimens These results were established by several landmark studies during 1970s and 1980s. They demonstrated the efficacy of adjuvant chemotherapy and later neoadjuvant chemotherapy.
Considering this, standard chemotherapy for adult patients frequently does not include high-dose methotrexate. A comprehension of the outcomes of these patients without methotrexate is important to understand the disease scenario of this selected population and to guide the development of new treatment strategies with acceptable tolerance. The present study aimed to evaluate the outcomes of adult patients with high-grade localized osteosarcoma treated with chemotherapy regimens without methotrexate.
Due to the clinical importance of the subject, we believe that our work would be of interest for ESMO Open readers.
With the submission of this letter I would like to undertake that the above-mentioned manuscript has not been published, accepted for publication and neither is under editorial review for publication elsewhere.
All authors contributed to conception, drafting the article and final approval of the version being submitted.
Sincerely,
The authors.
Corresponding author : Veridiana Pires de Camargo, MD
Instituto do Cancer do Estado de Sao Paulo

